# Central nervous system immune response in postinfectious hydrocephalus

**DOI:** 10.1101/2020.07.30.20165332

**Authors:** Albert M. Isaacs, Sarah U. Morton, Mercedeh Movassagh, Qiang Zhang, Christine Hehnly, Lijun Zhang, Diego M. Morales, Shamim A. Sinnar, Jessica E Ericson, Edith Kabachelor, Peter Ssenyonga, Justin Onen, Ronnie Mulondo, Mady Hornig, Benjamin C. Warf, James R. Broach, R Reid Townsend, David D. Limbrick, Joseph N. Paulson, Steven J. Schiff

**Affiliations:** Department of Neuroscience, Washington University School of Medicine; Department of Clinical Neurosciences, University of Calgary; Division of Newborn Medicine, Boston Children’s Hospital; Department of Pediatrics, Harvard Medical School; Department of Biostatistics, Harvard T.H. Chan School of Public Health; Department of Medicine, Washington University School of Medicine; Institute for Personalized Medicine, Pennsylvania State University; Department of Biochemistry and Molecular Biology, Pennsylvania State University; Department of Neurosurgery, Washington University School of Medicine; Center for Neural Engineering, Pennsylvania State University; Department of Medicine, Pennsylvania State University College of Medicine; Department of Pediatrics, Pennsylvania State College of Medicine; CURE Children’s Hospital of Uganda; Department of Epidemiology, Columbia University Mailman School of Public Health; Department of Neurosurgery, Harvard Medical School; Department of Biostatistics, Product Development, Genentech Inc; Department of Engineering Science and Mechanics, Pennsylvania State University; The Center for Infectious Disease Dynamics, Pennsylvania State University; Department of Neurosurgery, Pennsylvania State University College of Medicine; Department of Physics, Pennsylvania State University

## Abstract

Inflammation following neonatal infection is a dominant cause of childhood hydrocephalus in the developing world. Understanding this complex inflammatory response is critical for the development of preventive therapies. In 100 African hydrocephalic infants ≤3 months of age, with and without a history of infection, we elucidated the biological pathways that account for this inflammatory response. We integrated proteomics and RNA sequencing in cerebrospinal fluid, identifying gene pathways involving neutrophil, interleukin (4, 12, and 13) and interferon activity associated with this condition. These findings are required to develop strategies to reduce the risk of hydrocephalus during treatment of infection.

## INTRODUCTION

Hydrocephalus is a serious brain disorder in children^1,2^ and the most common indication for pediatric neurosurgery worldwide^3^. The most frequent global antecedent of hydrocephalus is infection such as neonatal or infant sepsis^4^. Postinfectious hydrocephalus (PIH) accounts for up to 60%^5^ of the nearly 400,000 infants who develop hydrocephalus each year, principally in low- and middle-income countries^2^. Despite recent clinical efforts to optimize treatment^6^, PIH remains a leading cause of neurological morbidity and mortality worldwide^7,8^. Strategies to prevent PIH have been thwarted for two principal reasons. First, the key pathogens responsible for the underlying infectious episodes that precede PIH are often not identified, and thus treatment of the underlying infections is not optimized^9^. Second, we have not identified the specific underlying inflammatory milieu that leads to hydrocephalus following infection^10,11^. Unfortunately, even children who undergo surgical treatment for hydrocephalus early in life suffer very poor long-term outcomes^7,12^. Therefore, major advances in the health of these children will require preventing infection by targeting both the underlying pathogens and their routes of infection^13-15^, as well as treating infections with adjunctive therapies that can reduce the likelihood of subsequently developing hydrocephalus.

A recent pan-microbial approach uncovered *Paenibacillus spp*. as a novel PIH pathogen associated with severe ventriculitis in a cohort of Ugandan children^16^. We hypothesize that the PIH host response is mediated by the activation of specific gene pathways, and that uncovering these pathways may identify therapeutic targets in hydrocephalus prevention.

To test this hypothesis, we performed high throughput proteomics and RNA sequencing (RNA-Seq) of cerebrospinal fluid (CSF) from 64 PIH infants and 36 non-postinfectious hydrocephalus (NPIH) infants to examine the immunopathogenesis of PIH ^16-18^. Integrating deep-scale proteomics (i.e., high gene coverage) with genomic and/or transcriptomic data, known as proteogenomic analysis^19,20^, permits the exploration of gene networks and pathways underlying disease. To our knowledge, this is the first proteogenomics study analyzing the interactions between PIH-associated proteome and transcriptome to better understand gene regulatory networks involved in PIH (Figure 1).

**Figure 1:**
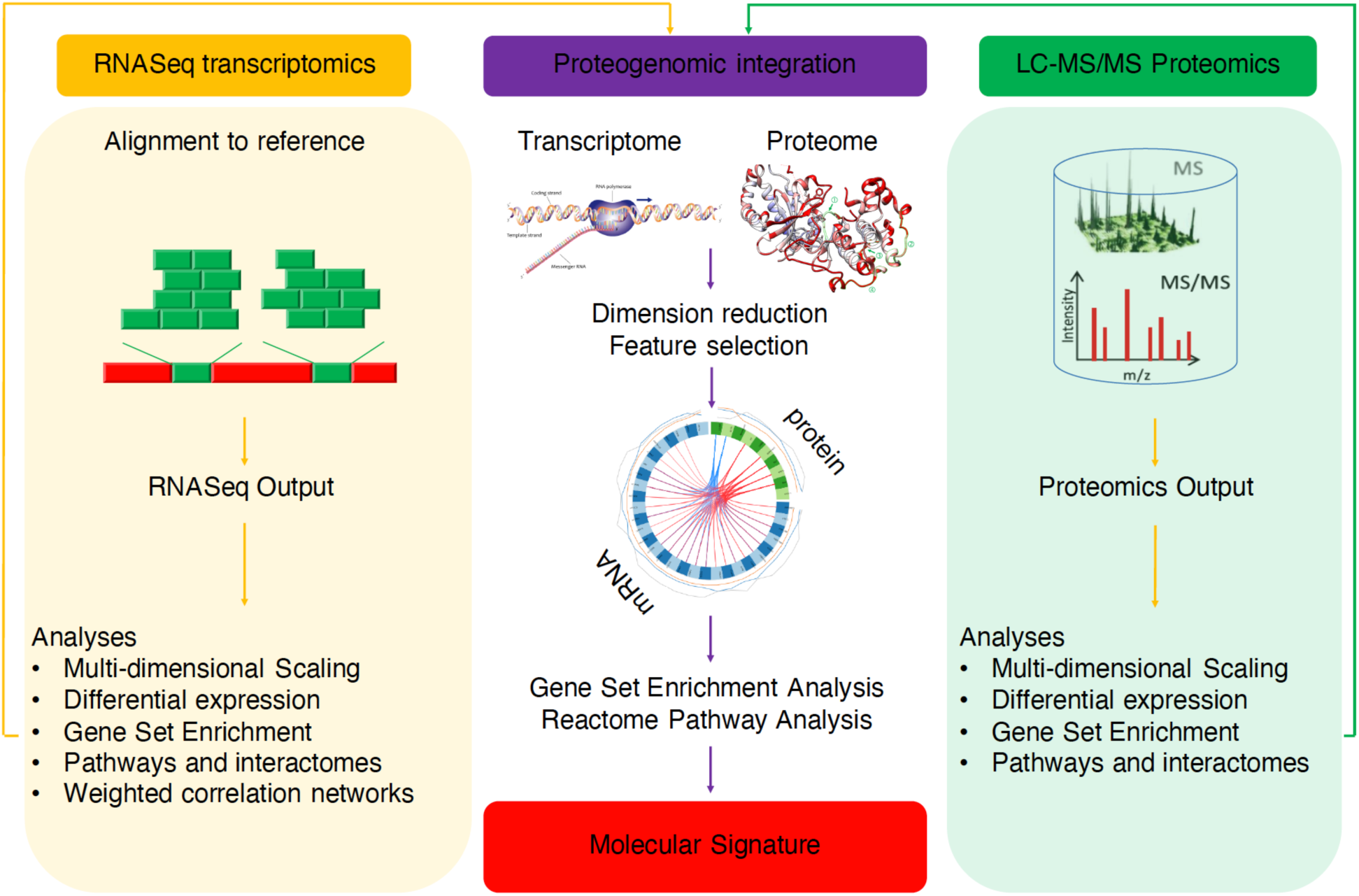
Schematic of proteogenomics experimental and data analysis workflow. Output data from concurrent proteomics and RNA-Seq on the same samples were preprocessed, normalized, batch-controlled and explored independently. Differentially expressed transcriptomic and proteomic data were integrated following dimension reduction and feature selection. Gene ontology enrichment was evaluated and common reactome pathways were visualized to identify prominent molecular pathways implicated in the pathophysiology of postinfectious hydrocephalus.

## RESULTS

### Patient characteristics

A total of 100 patients (64 PIH, 36 NPIH), all 3 months of age or less and with weight greater than 2.5 kg, were recruited. PIH infants had either a history of febrile illness and/or seizures preceding the onset of hydrocephalus, or alternative findings such as imaging and endoscopic results indicative of prior ventriculitis (septations, loculations, or deposits of debris within the ventricular system). None of the PIH infants had a history of hydrocephalus at birth. NPIH patients had findings of hydrocephalus that was of non-infectious origin on computed tomography (CT) scan or at endoscopy, including a structural cause (obstruction of the Aqueduct of Sylvius, Dandy-Walker cyst, or other congenital malformation) or evidence of hemorrhage (bloody CSF). Prior to CSF acquisition, none of the 100 patients had undergone surgery on the nervous system (reservoirs, shunt, third ventriculostomy, or myelomeningocele closure), or had evidence of communication between the nervous system and skin such as meningocele, encephalocele, dermal sinus tract, or fistula.

Sterile CSF samples acquired directly from the cerebral ventricles of all patients during surgical treatment of hydrocephalus (shunt insertion or endoscopic treatment) in Uganda were placed into cryotubes with DNA/RNA preservative (DNA RNA Shield, Zymo Corp). Samples were then frozen in either liquid nitrogen or at -80°C and shipped cryogenically to the USA for further processing. Previous 16S rRNA sequencing had recovered *Paenibacillus spp*. from 59% of the PIH infants in this study, of which 8 and 27 patients also had Human Herpesvirus 5 (cytomegalovirus, CMV) in the CSF or blood^16^, respectively. These previous findings directed stratification of our patients into “Paeni-positive” or “Paeni-negative,” “CMV-CSF-positive” or “CMV-CSF-negative,” and “CMV-serum-positive” or “CMV-serum-negative.” Comparison of demographic and clinical attributes between Paeni-positive and Paeni-negative patients showed that Paeni-positive patients had higher white blood cell (WBC) counts in peripheral blood (11.9 vs 9.3 × 10^3^/µL, p=0.002) and CSF (71.6 vs 5.0 × 10^3^/µL, p<.001), and lower blood hemoglobin levels (10.8g/dL vs 12.0 g/dL, p=0.039) than those of Paeni-negative patients. There were no significant differences in age or gender between the groups (Table 1).

**Table 1:**
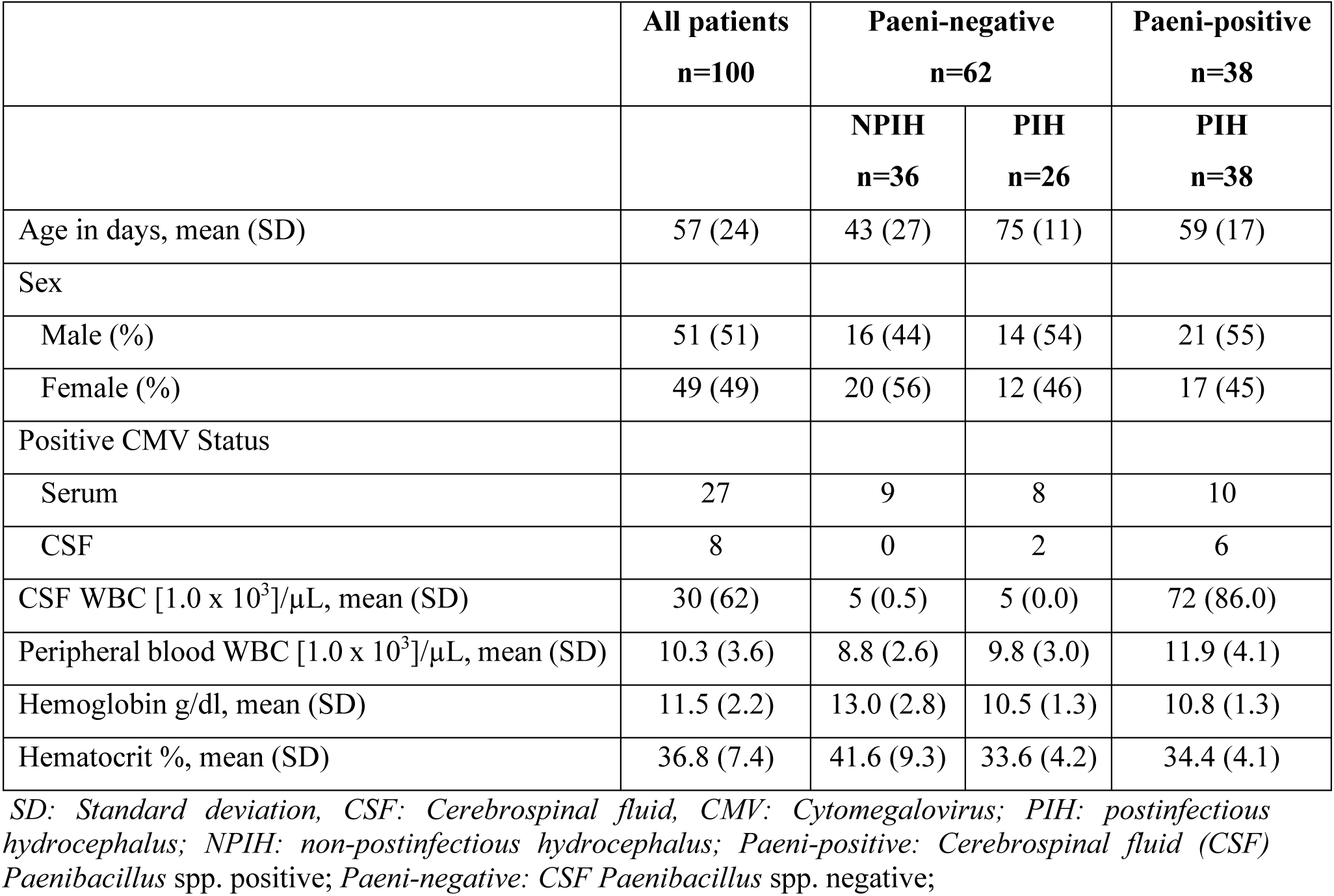
Demographics and clinical characteristics of patients based on *Paenibacillus* spp. status

### Protein expression

Tandem mass spectrometry (MS/MS) spectra obtained from a standard ten-channel peptide mass tagging (TMT-10) system and compared with the current UniRef^®^ database yielded quantitative proteomic data^17,21-23^. Median values of peptide intensities were assigned to unique proteins and were used to infer relative protein abundances. Peptide identifications that could be assigned to more than one protein were excluded from protein quantification. Batch effects were corrected, and intensity values were quantile normalized. Multi-dimensional scaling (MDS) analysis demonstrated the clustering of PIH and NPIH infants and distinguished Paeni-positive from Paeni-negative patients (Figure 2A). Of 616 proteins identified, 292 were differentially expressed based on *Paenibacillus spp*. status: 144 and 148 proteins were upregulated or downregulated in the Paeni-positive group, respectively (Figure 2B). Gene set enrichment analysis of the differentially expressed proteins identified the predominant enriched functions were involved in neuroinflammation, particularly neutrophil-mediated inflammation, and negative regulation of proteolysis and peptidase activity, and modulation of extracellular matrix and structure (Figure 2C).

**Figure 2:**
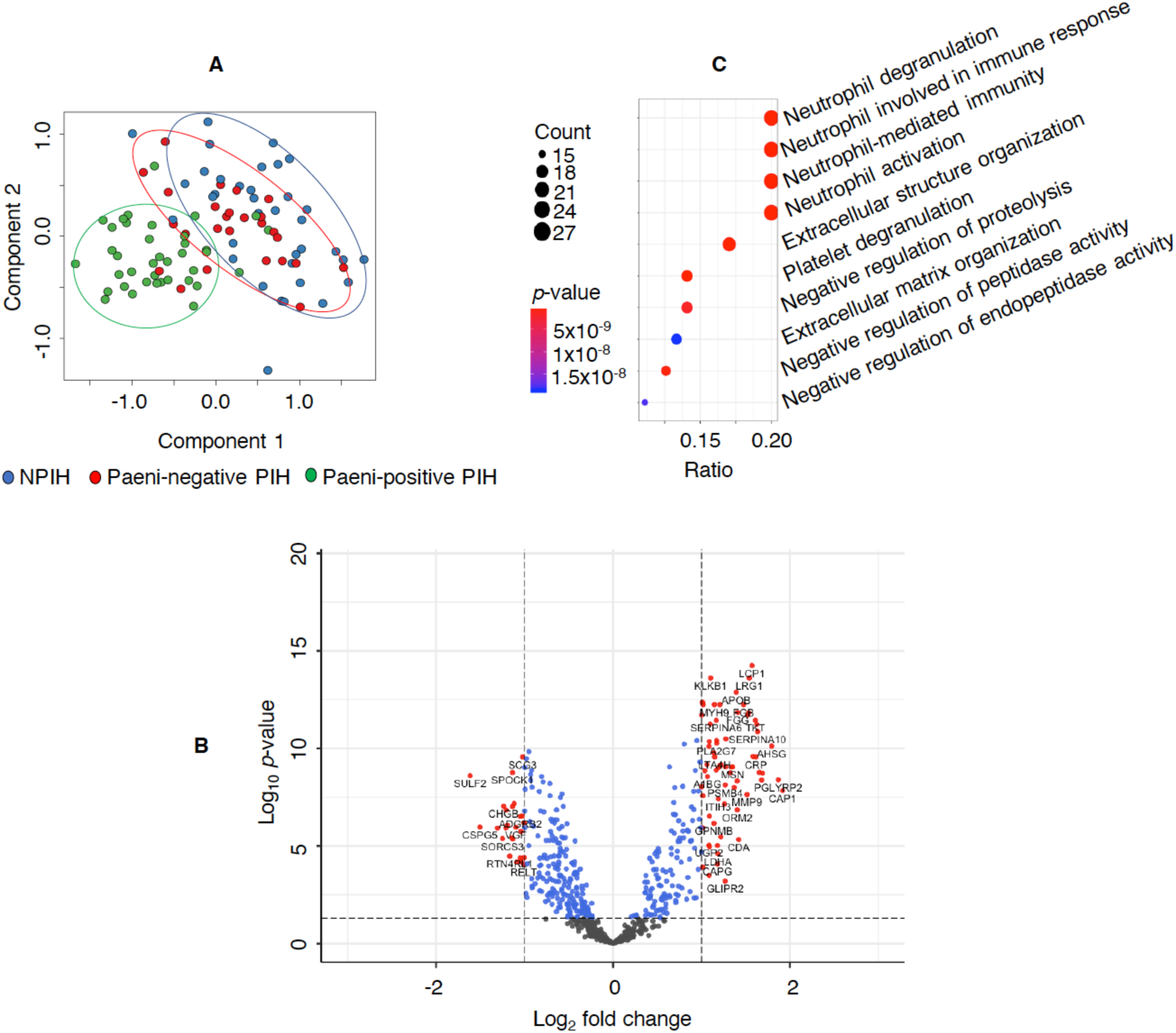
Proteomic profile of infants with postinfectious hydrocephalus (PIH) and those without (NPIH), based on 16s rRNA-determined *Paenibacillus spp*. status. Panel A is a Multi-dimensional Scaling of normalized protein abundances demonstrating clustering of *Paenibacillus spp*. positive (Paeni-positive) infants from the remaining groups. The abscissa and ordinate of the scatter plot of individual participants represent the first and second components, respectively. Each oval (not drawn to scale) encircles majority of patients belonging to the color-matched group, with blue as infants in the NPIH group, red as Paeni-negative PIH infants, and green as Paeni-positive infants. Panel B is a volcano plot demonstrating differential expression of genes between Paeni-positive and Paeni-negative infants. Adjustment for cytomegalovirus status did not change the differential expressions between groups. The vertical lines crossing the positive and negative abscissae demarcate fold changes of 1 and -1 respectively, and the horizontal dashed line crosses the ordinate at the alpha significance level of 0.05. Each point represents a differentially expressed protein and those with an absolute fold change greater than 1 that met the significance level (red points) were selected for gene set enrichment analyses. Genes with enrichment below the statistically significant threshold are displayed in grey. Panel C is a Gene ontology analyses of differentially expressed proteins of infants based on 16s rRNA-determined *Paenibacillus* spp. status. There was enrichment for functions associated with neuroinflammation, extracellular matrix structure and cell-cell adhesion among Paeni-positive infants compared to Paeni-negative infants. The abscissae (ratio) of the dot plots correspond to the number of proteins per total number of proteins and the size of each circle reflects the relative number of proteins expressed that are enriched for the corresponding function (ordinates).

### RNA Expression

Gene read counts (25 million reads per sample) were aggregated from expression levels estimated from paired-end RNA-Seq data mapped to the human reference genome hg38 with STAR, and quantified with RESM^*24,25*^. Expression of at least 1 count per million in a minimum of 18 samples was required for inclusion in the analysis. MDS analysis demonstrated the clustering of PIH and NPIH infants (Figure 3A). Of 11,114 genes, 2,161 were differentially expressed, and the hierarchical clustering of differentially expressed genes distinguished Paeni-positive from Paeni-negative patients (Figure 3B). Gene ontology analysis of the differentially expressed genes based on *Paenibacillus spp*. status demonstrated enrichment for genes predominantly related to response to bacteria, host immune regulation, and cell motility, migration, and adhesion (Figure 3C).

**Figure 3:**
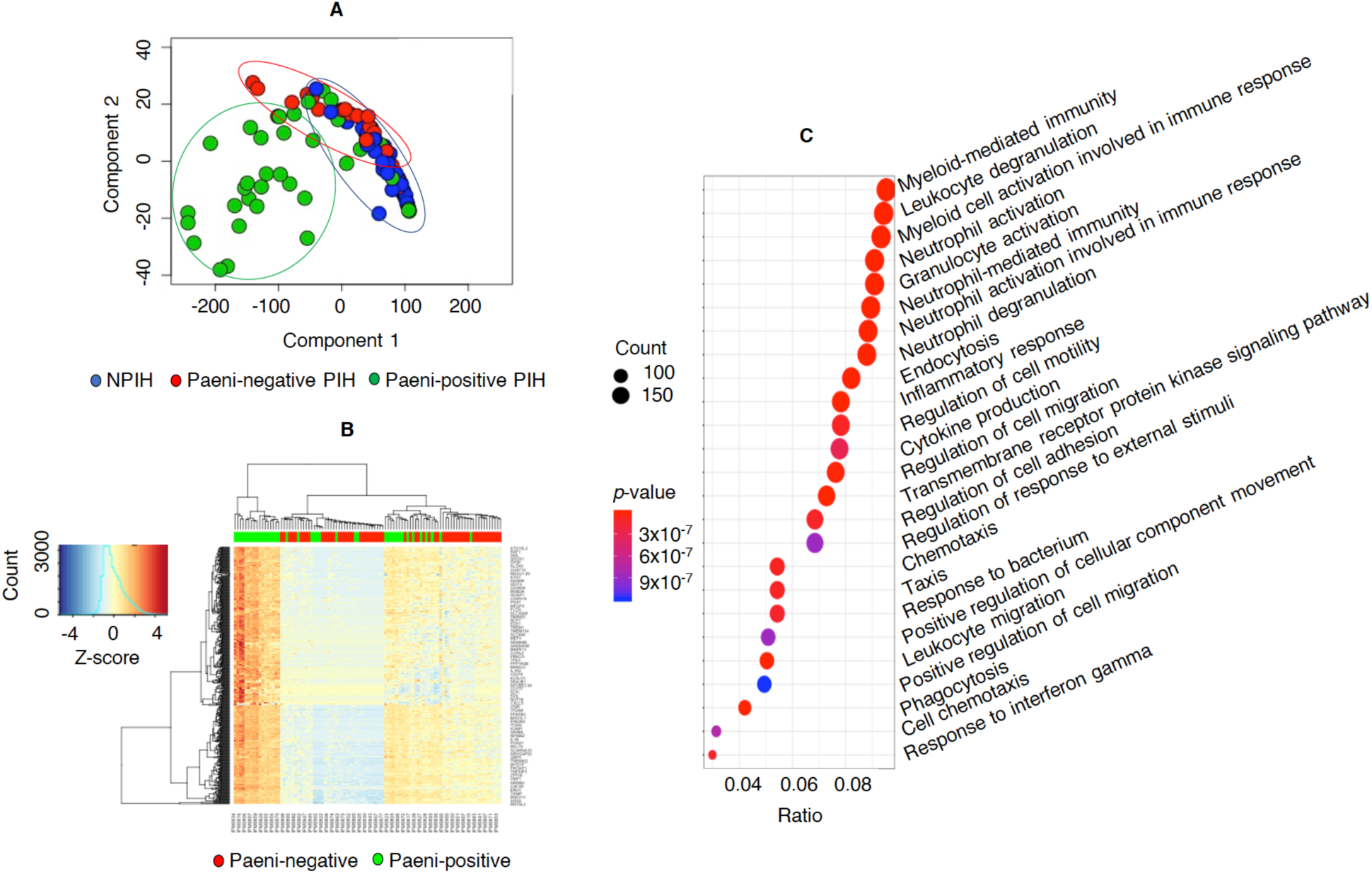
Transcriptomic profile of infants with postinfectious hydrocephalus (PIH) and those without (NPIH), based on 16s rRNA-determined *Paenibacillus* spp. status. Panel A is a Multi-dimensional Scaling of normalized gene expression demonstrating clustering of *Paenibacillus spp*. positive (Paeni-positive) infants from the remaining groups. The abscissa and ordinate of the scatter plot of individual participants represent the first and second components, respectively. Each oval (not drawn to scale) encircles majority of patients belonging to the color-matched group, with blue as infants in the NPIH group, red as Paeni-negative PIH infants, and green as Paeni-positive infants. Panel B is a Heatmap of the 500 most differentially expressed genes between the Paeni-positive and Paeni-negative infants, with the dendrogram demonstrating hierarchical clustering on Euclidean distance of gene identified in infants based on Paenibacillus spp. status. Panel C is a Gene ontology analyses of differentially expressed genes of infants based on 16s rRNA-determined *Paenibacillus* spp. status. There was enrichment for functions associated with neuroinflammation, extracellular matrix structure and cell-cell adhesion among Paeni-positive infants compared to Paeni-negative infants. The abscissae (ratio) of the dot plots correspond to the number of proteins per total number of proteins and the size of each circle reflects the relative number of proteins expressed that are enriched for the corresponding function (ordinates).

### Role of CMV co-infection

Adjusting for CMV status at the transcriptome level did not significantly alter the results of the gene ontology enrichment analysis based on *Paenibacillus spp*. positivity. Sixty-four genes were differentially expressed based on CMV status, and those genes were enriched for functions related to host response to virus (Figure 4A). Principal component analysis (PCA) using RNA abundance of all genes was not able to cluster samples by CMV status (Figure 4B). Adjusting for CMV status on proteomic data did not change the differentially abundant proteins (Figure 2C).

**Figure 4:**
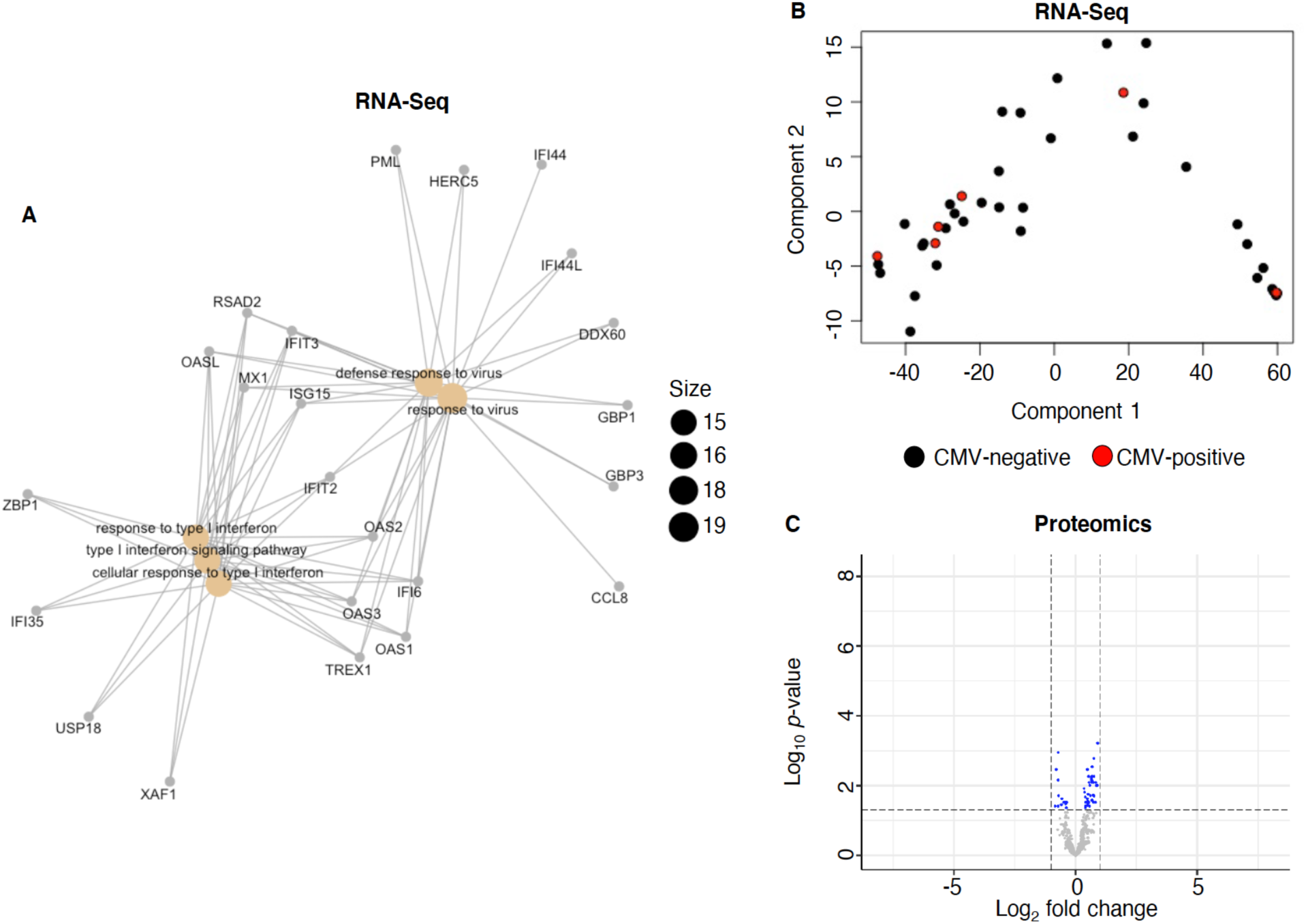
Role of CMV coinfection. Panel A is an interactome of the 64 genes that were differentially expressed based on CMV status, enriched for functions related to host response to virus. Panel B is a Principal Component Analysis plot demonstrating that using RNA abundance of all genes was not able to cluster samples by CMV status. The abscissa and ordinate of the scatter plot of individual participants represent the first and second components, respectively. Panel C is a volcano plot of demonstrating there no differentially expressed proteins based on CMV status alone (blue points) that had a fold change greater than 1 nor met statistical significance criteria. Genes with enrichment below the statistically significant threshold are displayed in grey. Differential expression of proteins based on *Paenibacillus* spp. status was similar with or without adjusting for CMV status (Figure 2).

### Correlation of clinical traits with gene expression

Network correlation analysis was employed to identify sets of genes that share a pattern of expression among patient RNA data. First, a subset of patients was identified using hierarchical clustering based on expression of genes that were differentially expressed between Paeni-positive and Paeni-negative patients. Then, using Weighted Correlation Network Analysis (WGCNA), 2 gene sets (Modules 1-2) were identified as having similar patterns of expression within this subset of 33 patients. These modules were assessed for network correlation with 4 clinical variables: CSF and peripheral blood WBC counts, *Paenibacillus spp*. status, and CSF CMV status (Figure 5A). Module 1 contained all but 27 of the 2,205 genes differentially expressed with respect to *Paenibacillus spp*. status. This module correlated positively with CSF cell count and negatively with *Paenibacillus spp*. status. Genes in Module 1 were enriched for functions related to host immune response (Figure 5B). Module 2 was positively correlated with *Paenibacillus spp*. status, but the 27 genes included in this module were not enriched for any gene ontologies, indicating no specific functional enrichment for the small number of genes within this module.

**Figure 5:**
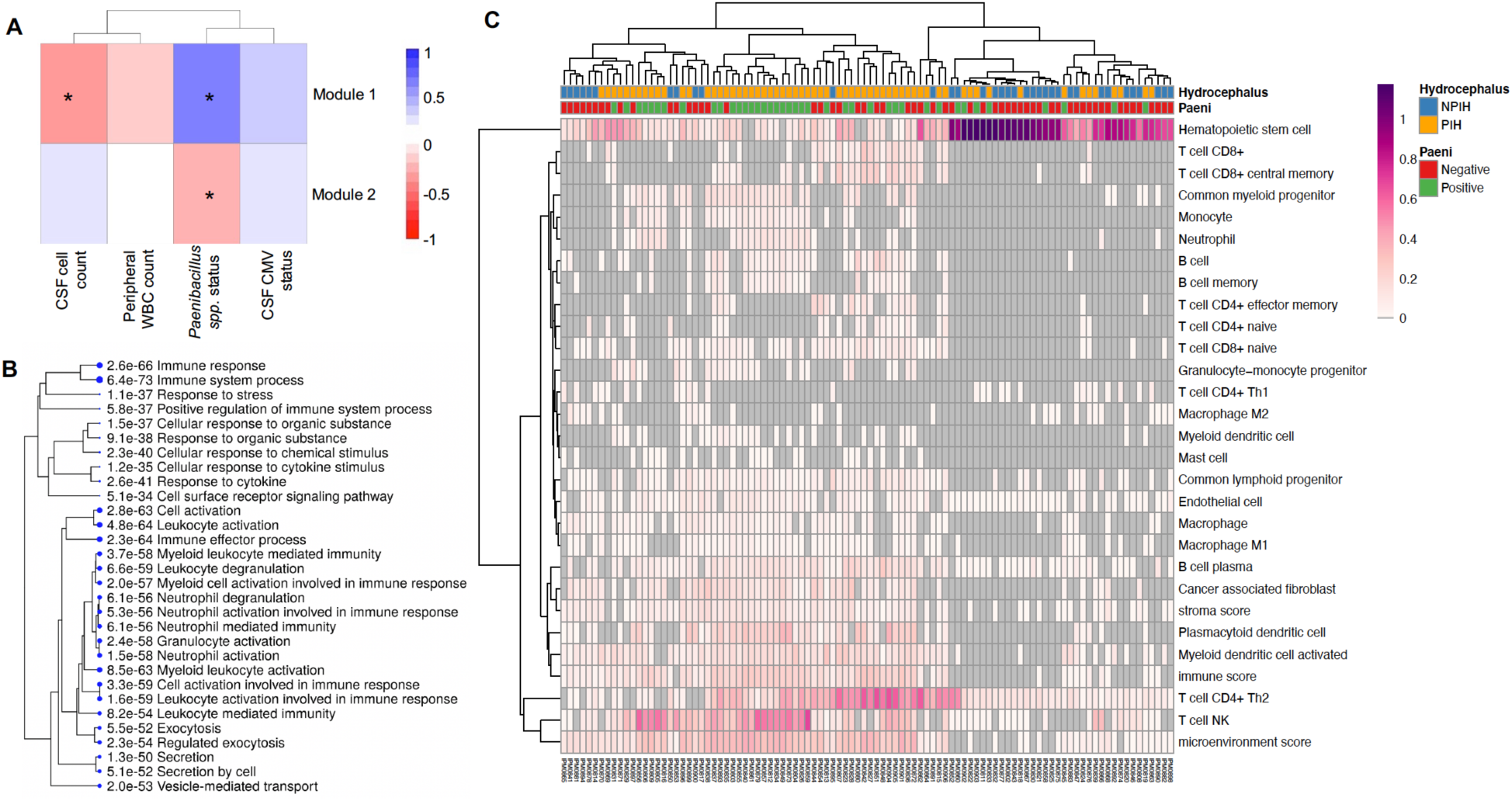
Weighted correlation network analysis (WGCNA) and single cell RNA deconvolution. Panel A demonstrates gene modules within RNA-Seq data for PIH and NPIH cohorts identified by WGCNA. Module 1 negatively correlated with CSF cell count and positively with *Paenibacillus spp*. status and was enriched for host immune responses, including leukocyte and neutrophil functions (Panel B). Module 2 was negatively correlated with *Paenibacillus spp*. status, but the associated genes had no specific functional enrichment. Panel C is deconvolution of bulk RNA data into immune cell populations expressed on a scale of 0-1 that calculated for each patient. There was hierarchical clustering of a mixture of NPIH and PIH samples with a hematologic predominance, and PIH-only samples with T-helper and NK cell populations.

### Marker genes of bulk RNA sequencing

Patient bulk RNA expression was analyzed for immune cell-type signatures using reference single-cell RNA sequencing data (Figure 5C). The *R* package xCell identifies cell type proportions within bulk RNA sequencing data based on enrichment of marker gene expression^26^. Upon hierarchical clustering, two groups were evident: a mixture of PIH and NPIH samples with a predominance of hematologic cells and PIH-only samples with T-helper and natural killer (NK) cell populations.

### Proteogenomic integration

Integration of proteomics and transcriptomics data was performed following dimension reduction and feature selection of differentially expressed proteins and genes. Unsupervised gene ontology analyses of the concatenated data recapitulated enrichment for functions related to the immune system, metabolism, response to oxidative stress, cell-cell junction interactions, and extracellular matrix structure in association with peptidase activity (Figure 6A).

**Figure 6:**
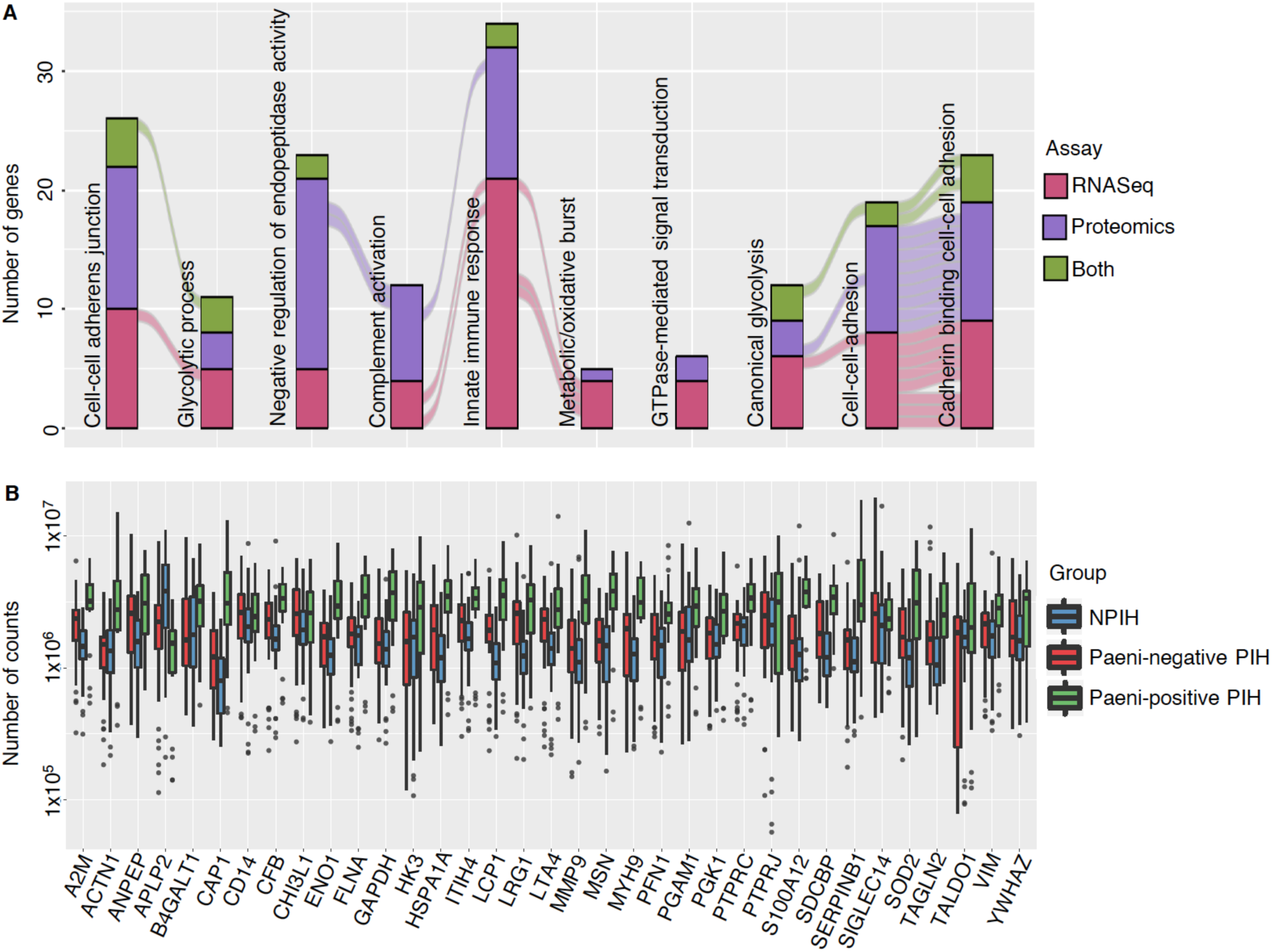
Proteogenomic integration of proteins and genes expressed in RNA-seq and/or proteomics among postinfectious and non-postinfectious hydrocephalus infants stratified by cerebrospinal fluid 16s rRNA *Paenibacillus* spp. status. Panel A is an Alluvial plot demonstrating the most prominent Gene Ontological (GO) functions and interactions for PIH pathophysiology between *Paenibacillus* spp. positive and *Paenibacillus* spp. negative infants. Each ontological clustering occupies a column in the diagram and is horizontally connected to preceding and succeeding significance clustering by stream fields, representing similar gene involvement. Each stacked bar is color-coded based on the assay being assessed, with red representing RNA-Seq data, purple for proteomics and green for genes common to both RNA-Seq and proteomics. The ordinate shows the number of genes represented in each cluster.

Panel B is a Box and Whisker plot of the 33 genes that were differentially expressed based on *Paenibacillus spp*. status. Counts (ordinate) of each gene (abscissa) are shown for each group, with blue representing NPIH, red for Paeni-negative PIH and green for Paeni-positive PIH infants.

There were 33 genes detected by both proteomics and RNA-Seq as significant; Paeni-positive PIH infants had consistently differing counts of those markers than did the Paeni-negative and NPIH infants (Figure 6B). Pathway enrichment analysis of those genes demonstrated a predominance of functions associated with the immune system, particularly those involved with interleukin (IL)-4, IL-12, IL-13, interferon, and neutrophil activity, as well as those relating to Janus kinase/signal transducers and activators of transcription (Jak/STAT) pathway. In addition, there was enrichment for processes involving response to platelet-activating factors and host recognition of microbes including antigen presentation and Class I MHC antigen processing (Figure 7).

**Figure 7:**
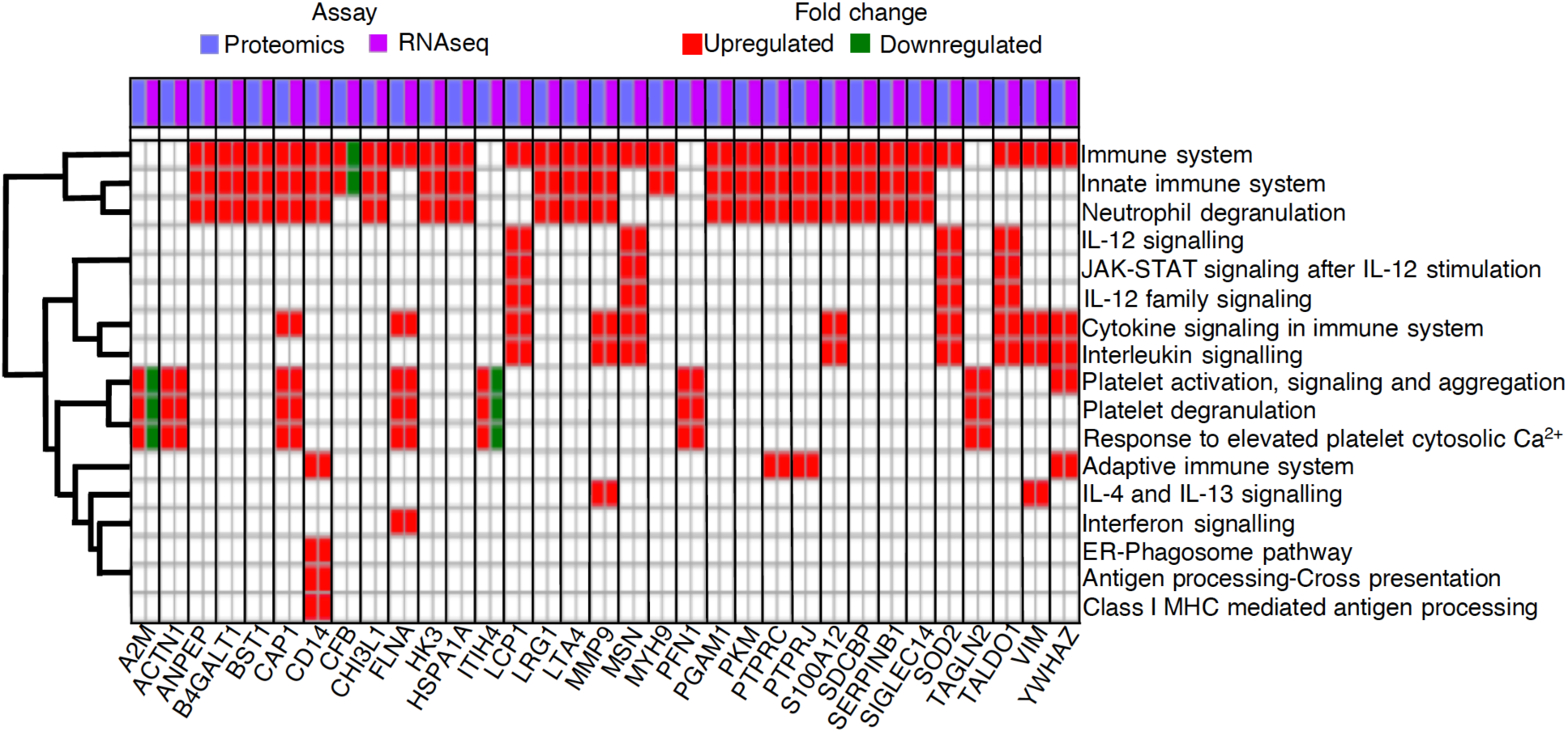
Pathway analysis of 33 genes that were differentially expressed in both RNA-seq and proteomics among postinfectious and non-postinfectious hydrocephalus infants stratified by cerebrospinal fluid 16s rRNA *Paenibacillus* spp. status. Corresponding pairs of upregulated (red boxes) and downregulated (green boxes) proteins (blue columns) and genes (purple columns) in the *Paenibacillus* spp. positive group that were identified with proteomics and RNA-Seq respectively are listed on the abscissa. The 33 common genes demonstrated predominant involvement of the immune system, particularly the innate system and those associated with neutrophil-mediated activity, interleukins, interferon and the Janus kinase/signal transducers and activators of transcription (JAK-STAT) pathway (ordinate). Differential expression was defined as log2 fold change of >1 or <-1 at an alpha significance level of 0.05.

## DISCUSSION

PIH is a debilitating neurological sequela of neonatal sepsis^4,7,8^. We identified critical gene products and pathways involved in the inflammatory mechanisms early in PIH. To our knowledge this study is the first to utilize proteogenomics to characterize gene regulatory networks involved in any form of hydrocephalus. These findings inform the development of adjunctive strategies for neonatal sepsis treatments to prevent hydrocephalus.

Large scale proteomics and genomics studies have been instrumental in the discovery of important biomarkers and have provided insights into the pathophysiology of central nervous system disease. CSF proteomics has previously identified clinically useful protein biomarkers such as amyloid precursor protein and cell adhesion molecules in PHH^17,27^. DNA or RNA sequencing from CSF has been widely used to study infectious diseases^28,29^. Because disease pathogenesis involves the combinatorial interaction of elements within the proteome, transcriptome, and environmental factors^30,31^, integrative methods that combine proteomic and genomic data into a network model are essential for thoroughly understanding human disease^32^. Here we utilized clustering, pathway analysis, and gene ontology enrichment methodologies at the RNA and protein levels to unravel the underlying mechanisms involved in the host response to PIH.

### Summary of findings

In this study, genes with differential RNA or protein abundance among PIH patients were enriched for functions related to neuroinflammation, reaction to oxidative stress, cell-cell junction structure, and extracellular matrix organization. Combining proteomics and RNA-Seq approaches narrowed the spectrum of host responses to predominantly those involved with the innate immune system, including neutrophil activity, signaling via IL-4, IL-12, IL-13, and interferon, and Jak/STAT pathways, in addition to platelet response/activating factors. Furthermore, there was enrichment for factors involved with microbe recognition such as Class I MHC antigen-presenting complex. In light of the recent confluence of evidence suggesting that dysregulated neuroinflammation and related ependymal scarring and aqueductal obstruction propagate inflammatory hydrocephalus (including PIH and PHH)^4,11^, the pathways we identified are potential targets for adjunctive treatments to reduce the risk of neuroinflammation and hydrocephalus following neonatal sepsis.

### Host immune responses in PIH

In addition to ventriculomegaly (enlargement of the cerebral ventricles), the PIH phenotype is characterized by CSF fluid loculations, debris within fluid spaces, ectopic calcification, and brain abscesses^8^, suggesting that severe inflammation occurs locally during the antecedent neonatal sepsis. Typically, once a pathogen breaches a host’s epithelial barriers, immune mechanisms are activated. Although there is a rapid progression of immunologic competence after birth, the innate immune system is the primary active defense for infants less than 3 months of age, since they lack the antigenic experience that informs acquired immunity. Consequently, the neonatal host immune response involves antigen-independent immune components such as neutrophils, phagocytes, NK cells and antigen-presenting cells^33,34 35-37^. Our RNA-Seq immune cell signature analyses support the role of NK cells in the host response of PIH patients. The Toll-like Receptor (TLR) immune system pathway is commonly implicated in post-inflammatory (including infectious or hemorrhagic) hydrocephalus^11,38-40^; however, the cytokine and interferon induced Jak/STAT pathway is considered to be the most efficient form of innate immunity, especially with intracellular pathogens^37,41^. Protein mutations in the Jak/STAT pathway have been implicated in poor inflammation mediation^37,41^. For example, impaired Jak function in severe combined immunodeficiency is associated with susceptibility to infections due to the absence of NK, B, or T cells^42,43^. At the protein and RNA levels, we observed differential expression of factors associated with interleukins (IL-4, IL-12 and IL-13) and interferons activity, both of which are obligate mediators of the Jak/STAT pathway^37,41^. Upregulation of MHC Class I in our cohort could be an indication of presentation to cytotoxic CD8+ cells and IL-12 activity^44^.

### Inflammation and barrier integrity in PIH

The activated immune pathways found in PIH support the perspective that the neonatal immune response to toxins generated by infection or hemorrhage inadvertently leads to ependymal gliosis or denudation, scarring of CSF conduits, and disruption of CSF physiology to cause hydrocephalus. Given that infants presenting with PIH are often remote from the original infection and clinically well for surgery, persistent inflammation in their CSF may be pathological rather than beneficially adaptive^11^. While the host-immune response is necessary for clearing microbial pathogens and fighting infections, prolonged or overstimulated immune activation may be detrimental to the host^45^. In PIH, it is possible that the same host immune activity that clears pathogens also causes hydrocephalus by damaging local tissues. Our finding of a differential abundance of RNA and proteins involved in cell-cell integrity pathways is consistent with recent evidence from both experimental^46-51^ and clinical^46,52,53^ studies demonstrating that host immune response is associated with ependymal cell-cell junction protein disruption as a critical pathogenetic mechanism of hydrocephalus. We also observed differential abundance of RNA and proteins associated with platelet-activating factors and response to reactive oxidative species. Elevations in platelet-activating factors have been associated with compromise of the blood brain barrier^54^, and their signaling pathways are an important link between inflammatory and thrombotic processes, including in sepsis^55^. Neutrophil-generated reactive oxidative species have also been implicated in disruptive blood-brain barrier changes during sepsis^56^. During an acute infection there is a high rate of oxidative metabolism and response to reactive oxidative species such as hydrogen peroxide, superoxide radicals and nitric oxide^57^, which typically sets off a cascade of cellular excitotoxicity and secondary brain injury that impairs brain oxygenation and perpetuates cerebral vascular dysfunction and ischemia^58^. Such vascular pathology might be associated with our finding of a differential expression of factors involved in platelet activity including PFN1, ITIH4 and A2M. The blood-brain barrier disruption may herald the extravasation of microbes and inflammatory mediators from the systemic compartments into the ventricles and other intracranial compartments.

### Potential pathologic overlap with NPIH

Inflammation is a component of other forms of hydrocephalus^38,59^, which informed our decision to use non-infectious hydrocephalic infants as controls; doing so allowed us to be able to detect molecular mechanisms that are more specific to PIH. In a recent review, Karimy et al. discussed evidence that PIH shares common host immune pathways with PHH^11^. In our cohort, we identified IL-12 signaling, which is typically involved in augmentation of CD8+ T cell cytotoxicity, as an important cytokine pathway underlying PIH. Interestingly, Habiyaremye et. al. recently reported PHH is associated with significantly elevated levels of IL-12, in addition to other cytokines including IL-1, IL-10, CCL-3 and CCL-13^59^. In an experimental model, TLR4-mediated host immune response was found to lead to choroid plexus hypersecretion of CSF^38^, and potential facilitation of PHH. Neuroinflammation has been linked to the ependymal barrier damage found in humans and several experimental models of hydrocephalus. In a human postmortem histological study, McAllister et. al. implicated glial fibrillary acidic protein over-expression, a non-specific surrogate marker of neuroinflammation and glial scarring in the diffuse ependymal damage they identified in PHH infants^60^. The ependymal damage was shown to result from cleavage of cell adhesion proteins^60,61^. Indeed, many forms of human congenital hydrocephalus result from genetic alterations of proteins involved in cell-cell junctional integrity including N-cadherin, connexin, and L1CAM^62^, that are critical for the differentiation of the ventricular and subventricular zones neural stem cells into mature ependyma^63^. In addition, a recent exome sequencing of 381 human congenital hydrocephalus infants identified a predominance of *de novo* mutations in genes associated with neural stem differentiation^64^. Experimentally, a series of studies on *hyh* mice^49,51,65-67^ that develop perinatal aqueductal stenosis also support the concept of a defect in cell junction complexes as an underlying cause of hydrocephalus. Collectively, these findings emphasize a critical association between host-immune response, cell-cell junction regulatory molecules and hydrocephalus, and raise the question as to whether modulation of the inflammatory signals common to PIH and NPIH might be beneficial.

### Role of immunomodulation for PIH prevention

This study supports the role of the immune response in PIH pathophysiology. Host immune response was the most predominant biological process among proteins and genes with differential abundance in PIH. However, it would be premature to suggest that our findings support the general use of anti-inflammatory or immunosuppressive agents during neonatal sepsis to mitigate the risk of developing hydrocephalus. There have been many attempts to use corticosteroids in treating infants with acute bacterial meningitis; but except for some notable exceptions such as *Hemophilus influenza*^68-72^ and pneumococcal meningitis^68- 73^, there have been many failures with some devastating outcomes^69,70,72-76^. The use of corticosteroids remains a controversial topic in perinatal medicine, and in particular, neonatal sepsis^77-80^. Our work however, points to novel intervention pathways and suggests that selective and targeted modulation of aspects of the immune response pathways may be a more successful approach to treating brain infections to prevent hydrocephalus. For example, in addition to pro-inflammatory immune factors, we found differential expression of markers associated with IL-4 and IL-13, which generally have anti-inflammatory effects. IL-4 and IL-13 can share a common receptor^81-83^ and typically work synergistically to decrease inflammation by counteracting the activity of pro-inflammatory cytokines such as IL-12^84,85^. IL-4 and IL-13 are also considered neuroprotective as they can induce death of microglial cells that mediate neuronal damage^86-88^. However, Park and colleagues^89,90^ report IL-4 and IL-13 potentiate oxidative stress-related injury to hippocampal neurons in experimental models. Thus, one hypothesis is that the balance of IL-12 and IL-4/IL-13 within the CSF contributes to risk of PIH, suggesting that their targeted modulation can be a potential therapeutic target. Further studies are needed to examine the role of cytokine/chemokine modulators in neonatal sepsis to prevent hydrocephalus.

### Viral co-infection in PIH

Identifying CMV in the CSF of a subset of infants in our cohort^16^ raises the important question of whether the host-immune response we observed was, at least in part, attributable to CMV. While there were differentially expressed transcripts detected in CSF and/or serum consistent with the host response to central nervous system viral infection in CMV-positive infants, we did not detect any functional proteins associated with a response to CMV, nor was there any effect of CMV status on the differential expression of host-immune markers at the protein level. The inability to detect a CMV-related host immune response at the protein level may be attributed to our small sample size (8/100 and 27/100) of CMV-CSF-positive and CMV-serum-positive infants, or the ability of the virus to modulate protein synthesis in the host^91^. Alternatively, CSF CMV status may underestimate CMV burden within the cohort, as intracellular CMV may be difficult to detect in CSF samples with low cell counts. Another possibility is that because CMV tends to cycle through latent or non-lytic and active states in affected individuals^92^, sampling CSF at any single point in time may not capture an active state. Thus, it remains unknown whether CMV was a risk factor for developing PIH, whether CMV can be implicated in the persistent host-immune response we observed, or whether the presence of this virus will have long-term effects in these infants.

## Limitations

Limitations to this study include small cohort size for subset analyses such as CMV positive participants, low RNA abundance in NPIH samples, and reduced availability of clinical covariates to assess potential correlations with gene abundance patterns. Furthermore, the majority of activated gene networks we identified in this cohort were pathogen-stratified based on *Paenibacillus spp*. status, possibly limiting generalizability for other pathogens. However, the concentrated, high incidence of PIH in Sub-Saharan Africa makes our findings directly relevant and applicable to PIH not just in globally under-resourced areas but also as it occurs in other developed regions. Nevertheless, important future work will test the generalizability of host response observed here to PIH related to other pathogens.

## CONCLUSIONS

Inflammation following neonatal infection is a dominant cause of childhood hydrocephalus in the developing world. Proteome and transcript expression analysis identified gene pathways involving neutrophil, interleukin (4, 12, and 13) and interferon activity critically associated with PIH in African infants. These findings enable the development of preventive hydrocephalus risk reduction strategies.

## Data Availability

All relevant data for this manuscript have been included in the methodology and results

